# The Impact of Peripartum Floor Lockdowns on Newborn Safety

**DOI:** 10.1101/2023.04.28.23289274

**Authors:** Barbara Neshek, David Ray Bryant, Justin Hruska, Deepak Gupta

## Abstract

**Background:** For the safety of newborns, infrastructural lockdown of peripartum floors may be imperative.

**Materials and Methods:** Cumulative data with no identifiers at all was collected from Midas Incident Reporting System reports and Security Logs reports for comparing incidence of newborn abductions four year-period (2015-2018) before the institution of lockdown with four year-period (2019-2022) since the institution of lockdown.

**Results:** During the entire eight years, there was not a single incident of newborn abduction reported in Midas Incident Reporting System and Security Logs.

**Conclusion:** The absent incidence of newborn abductions before and since peripartum floors’ lockdown should not deter keeping safety protocols enhanced and potential loopholes closed thus preemptively treating lax human factors while protecting precious and vulnerable newborns during their stay in peripartum floors.

## Introduction

Working in lockdown units can be cumbersome and intimidating for healthcare workers [2-3] when ingress as well as egress out of lockdown units is dependent on well-functioning computerized scanners restricting security badge-based access to authorized healthcare workers considering that healthcare workers may have concerns about their own safety in terms of inability to egress lockdown units in case of inadvertently malfunctioning badge-based access during emergencies like fires and attacks. However, for the safety of patients, especially newborns, lockdown of peripartum floors may be imperative [4-6]. Peripartum floors at Hutzel Women’s Hospital, Detroit Medical Center (DMC) were turned into lockdown units in 2019 to promptly resolve and eliminate the incidence of newborn abductions. As four years have passed, now was the time to evaluate if incidence has changed.

## Materials and Methods

After institutional review board adjudging this project as non-human participant research, cumulative data about code pinks/purples [7-9] with no identifiers at all was collected from DMC Midas Incident Reporting System reports and DMC Security Logs reports for the following to compare four year-period before the institution of lockdown with four year-period since the institution of lockdown:

- Total number of code pinks in 2015-2018
- Total number of code purples in 2015-2018
- Total number of code pinks in 2019-2022
- Total number of code purples in 2019-2022
- How many infants (code pinks) were found back in 2015-2018?
- How many children (code purples) were found back in 2015-2018?
- How many infants (code pinks) were found back in 2019-2022?
- How many children (code purples) were found back in 2019-2022?
- Where were infants (code pinks) found back in 2015-2018?
- Where were children (code purples) found back in 2015-2018?
- Where were infants (code pinks) found back in 2019-2022?
- Where were children (code purples) found back in 2019-2022?

## Results

During the entire eight years, there was not a single incident of code pink or code purple reported in DMC Midas Incident Reporting System and in DMC Security Logs, thus making it impossible to quantify our peripartum floors’ lockdowns having effects if any in preventing newborn (and child) abductions from our peripartum floors.

## Discussion

Code Pink may have begun as a code for neonatal resuscitation [10] but it now universally codifies infant abduction especially newborn abduction in peripartum floors. Radiofrequency identification tags keep newborns safe within the confines of peripartum floors while tracking scanners promptly alarm whenever newborns are moved beyond the confines of peripartum floors unaccompanied by authorized caregivers [11-12]. However, restricted badge-based access for professional caregivers to ingress and egress peripartum floors makes those floors securer by allocating manageable resources to monitor badge-less ingress and egress of patient’s families and their visitors at the very few designated access points. Figures 1-2 are demonstrating that for the number of free access points to the unit reducing from nine to zero while keeping the two badge-based access to elevators same as before the change, the total number of access points itself has gone down from a total of eleven to four with one stairwell and six elevators closed to all access after the changes at the intrapartum floor as well as at the postpartum floor wherein key-based access to emergency elevator for emergency personnel use only may not be counted as access point to the unit. By limiting the number of designated badge-less access points just like any other critically important facility under strict and strong surveillance with restricted and monitored access of authorized personnel and people only [13-17], it becomes easier to prevent newborn abductions by potentially deterring potential abductors unless they have been criminally resourceful to illegally acquire the authorized badges providing them free access to peripartum floors. Moreover, like any other medical device or pharmaceutical drug, the radiofrequency identification tags can sometimes be in shortage and on backorders thus making the permanently changed infrastructure with restricted badge-access based peripartum floors lockdowns as a fail-safe mechanism for newborn safety. Additionally, presence of restricted badge-based access under strictly monitored movement surveillance may induce permanent change in the behaviors [18-27] of monitored personnel and people (providers and patients) by keeping them innately on their toes and always on vigil that even temporarily malfunctioning surveillance modes [28] may not increase the incidence of detectable and thus reportable incidents during Electronic Safety and Risk Management [29] via Midas Incident Reporting System.

**Figure 1:**
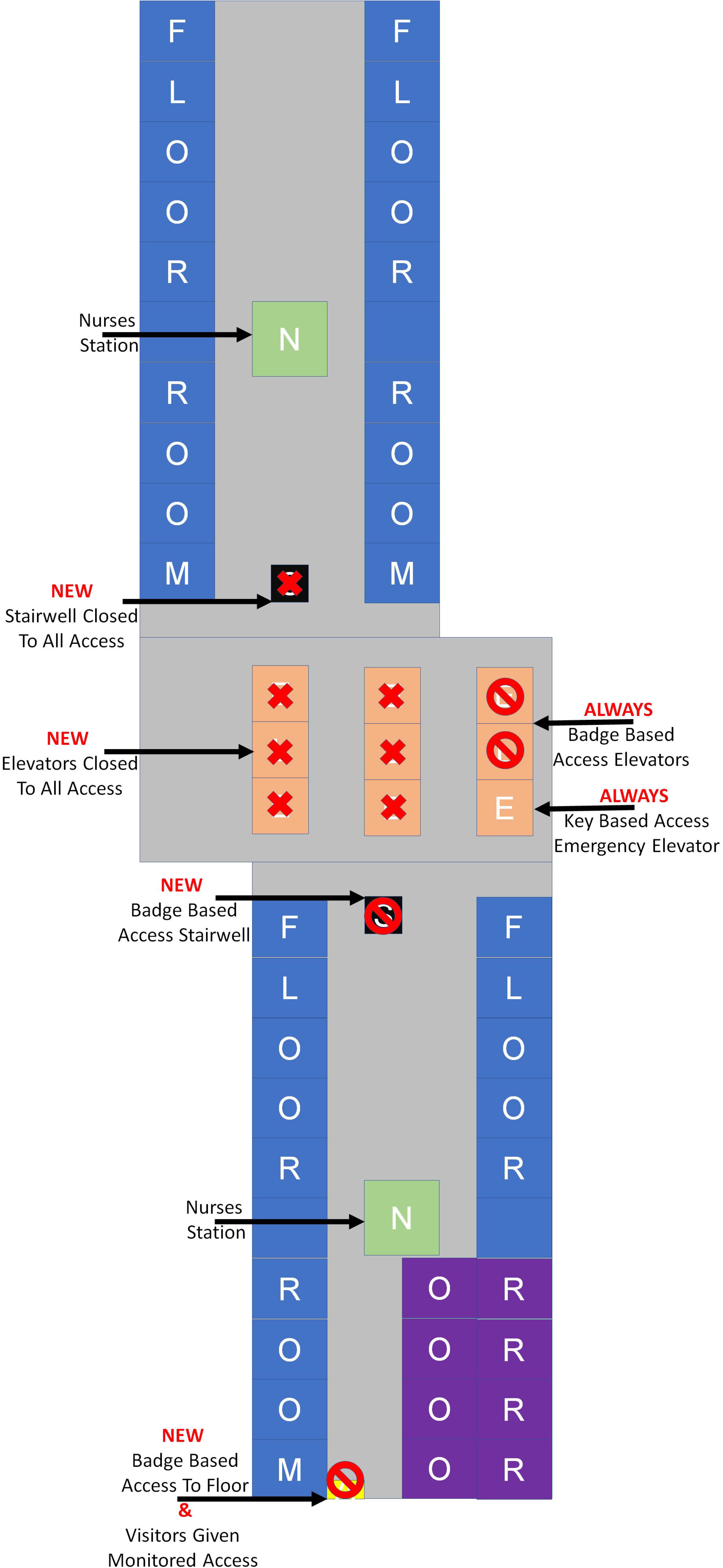
INTRAPARTUM FLOOR SCHEMATIC: SIMPLIFIED AND NON-EXACT LAYOUT SHOWING LABOR AND DELIVERY SUITE (FLOOR ROOMS) WITH OPERATING ROOMS (ORs) ALONG WITH BADGE BASED ACCESS AND NO ACCESS POINTS

**Figure 2:**
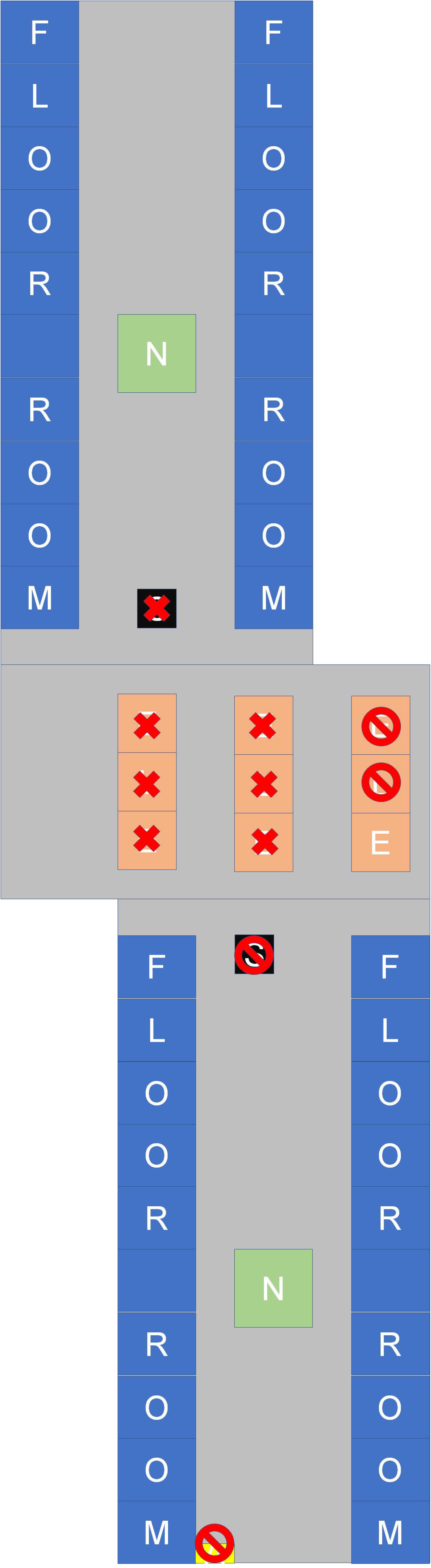
POSTPARTUM FLOOR SCHEMATIC: SIMPLIFIED AND NON-EXACT LAYOUT EXACTLY SAME AS INTRAPARTUM FLOOR SCHEMATIC EXCEPT FOR OPERATING ROOMS (ORs) WHICH ARE REPLACED WITH MORE FLOOR ROOMS

There were few limitations of our current study. Newborn abductions from healthcare institutions are already becoming rarer [30] thus making it difficult to gauge the effects of infrastructural changes to further newborn safety unless the study can be designed to investigate and analyze observations over a much longer period say over few decades rather than over few years. Considering that pre-crimes [31] in terms of pre-abductions can be quantified via the number of alarms detected per the planned unauthorized movements of newborns wearing radiofrequency identification tags beyond the confines of peripartum floors [32], it may be an interesting aspect to investigate in future studies because although enhanced security protocols like lockdown might have prevented pre-crimes in peripartum floors, such near-hits-near-misses could not be quantified during the current study.

## Conclusion

The absence of reported code pinks or purples before and since peripartum floors’ lockdown should not deter keeping safety protocols enhanced and potential loopholes closed thus preemptively treating lax human factors [33-35] while protecting precious and vulnerable newborns during their stay in peripartum floors.

## Supporting information

Non-Human Participant Reseach Per IRB

## Data Availability

All data produced in the present work are contained in the manuscript.

## Acknowledgement

The authors are indebted to the team of Lisa Katzman, Kathy King, Anne Chahine, and Marvin L Rutherford, at Detroit Medical Center (DMC), Detroit, Michigan, United States for running query in DMC Midas Incident Reporting System and DMC Security Logs for historical reports essential to complete the current study.

## References

1. The Impact of Peripartum Floor Lockdowns on Newborn Safety - Neshek - QuESST 2023 https://www.youtube.com/watch?v=9wLinaJnqbA

2. Joseph J. Weiss, MD Memorial Essay Contest Entry: Robohumans https://content.villagepress.com/DMN/4Q20/files/basic-html/page10.html

3. Joseph J. Weiss, MD Memorial Essay Contest Entry: Dynamic Lockdown https://pageturn.vpdemandcreationservices.com/DMN/3Q19/mobile/index.html#p=26

4. Security in Hospitals: How Hospitals Can Protect Infants. https://www.rft.com/infant-security-in-hospitals-how-hospitals-can-protect-infants/

5. Vincent JL. Infant hospital abduction: security measures to aid in prevention. MCN Am J Matern Child Nurs. 2009 May-Jun;34(3):179–83. doi: 10.1097/01.NMC.0000351706.81502.d0. PMID: 19550261.

6. Hiner J, Pyka J, Burks C, Pisegna L, Gador RA. Preventing infant abductions: an infant security program transitioned into an interdisciplinary model. J Perinat Neonatal Nurs. 2012 Jan-Mar;26(1):47–56. doi: 10.1097/JPN.0b013e31824003a2. PMID: 22293642.

7. CODE PINK. https://www.the-hospitalist.org/hospitalist/article/123116/code-pink

8. Code Pink system in hospital. https://expresshealthcaremanagement.blogspot.com/2018/10/code-pink-system-in-hospital.html

9. What is a Code Pink/Purple? https://lluh.org/patients-visitors/visitors/security-services/crime-information/what-code-pinkpurple

10. Koch J. Code pink: a system for neonatal resuscitation. JOGN Nurs. 1978 Sep-Oct;7(5):49–53. doi: 10.1111/j.1552-6909.1978.tb00774.x. PMID: 251751.

11. Hugs Infant Protection https://www.securitashealthcare.com/solutions/infant-protection

12. Hugs Infant Tag https://www.securitashealthcare.com/products/hugs-infant-tag

13. Why You Should Avoid Using a Fake Security Camera https://adssecurity.com/who-you-should-avoid-using-a-fake-security-camera/

14. Security Cameras Make Us Feel Safe, but Are They Worth the Invasion? https://www.nytimes.com/2022/11/02/technology/personaltech/security-cameras-surveillance-privacy.html

15. Do Security Cameras Deter Crime? https://www.adt.com/resources/do-surveillance-cameras-statistically-reduce-crime

16. Is There Empirical Evidence That Surveillance Cameras Reduce Crime? https://www.mtas.tennessee.edu/knowledgebase/there-empirical-evidence-surveillance-cameras-reduce-crime

17. The Watchers: Assaults on privacy in America https://www.harvardmagazine.com/2017/01/the-watchers

18. How People are Changing Their Own Behavior https://www.pewresearch.org/internet/2015/03/16/how-people-are-changing-their-own-behavior/

19. Does Technological Surveillance Change Behavior https://apps.dtic.mil/sti/citations/AD1062596

20. Jansen AM, Giebels E, van Rompay TJL, Junger M. The Influence of the Presentation of Camera Surveillance on Cheating and Pro-Social Behavior. Front Psychol. 2018 Oct 16;9:1937. doi: 10.3389/fpsyg.2018.01937. PMID: 30386277; PMCID: PMC6198084.

21. I See You: Do Surveillance Cameras Change People’s Behavior? https://nationalinterest.org/blog/buzz/i-see-you-do-surveillance-cameras-change-peoples-behavior-127762

22. How Fear of Government Surveillance Influences Our Behavior https://lithub.com/how-fear-of-government-surveillance-influences-our-behavior/

23. How surveillance changes behavior https://www.axios.com/2019/09/07/surveillance-changes-behavior

24. Social Behavior in Public Space: An Analysis of Behavioral Adaptations to CCTV https://www.ojp.gov/ncjrs/virtual-library/abstracts/social-behavior-public-space-analysis-behavioral-adaptations-cctv

25. How Being Filmed Changes Employee Behavior https://hbr.org/2014/09/how-being-filmed-changes-employee-behavior

26. What Constant Surveillance Does to Your Brain https://www.vice.com/en/article/pa5d9g/what-constant-surveillance-does-to-your-brain

27. Tim Cook expects our behavior to change when we feel ‘surveilled all the time’ by tech: ‘It changes society in a major way’ https://www.cnbc.com/2022/06/09/tim-cook-our-behavior-will-change-when-we-feel-surveilled-by-tech.html

28. McKinnon JF. Infant abduction: taking a new look at “false” alarms. J Healthc Prot Manage. 2008;24(1):87–90. PMID: 18409456.

29. Electronic Safety and Risk Management definition https://www.lawinsider.com/dictionary/electronic-safety-and-risk-management

30. Analysis of Infant Abduction Trends. https://www.ojp.gov/library/publications/analysis-infant-abduction-trends

31. Pre-crime and Policing in ‘Minority Report’ https://www.wondriumdaily.com/pre-crime-and-policing-in-minority-report/

32. Hugs® Infant Protection: SOLUTION OVERVIEW https://www.securitashealthcare.com/sites/securitashealthcare.com/files/media/2023-02/DOC-12-44404-AI_Hugs_Solution_Overview_2023.pdf

33. Webster KLW, Stikes R, Bunnell L, Gardner A, Petruska S. Application of Human Factors Methods to Ensure Appropriate Infant Identification and Abduction Prevention Within the Hospital Setting. J Perinat Neonatal Nurs. 2021 Jul-Sep 01;35(3):258–265. doi: 10.1097/JPN.0000000000000554. PMID: 34330138.

34. Human error, lax reporting and MCAS all contributed to fatal Boeing crash https://insights.globalspec.com/article/12909/human-error-lax-reporting-and-mcas-all-contributed-to-fatal-boeing-crash

35. The human factors in LAX safety https://www.latimes.com/opinion/la-oew-meshkati28-2008nov28-story.html

